# Inducing Body-Weight Supported Postural Perturbations during Gait and Balance Exercises to Improve Balance after Stroke – A Pilot Study

**DOI:** 10.1101/2021.06.11.21257723

**Authors:** Amanda Meyer, Erica Cutler, Jill Hellstrand, Emily Meise, Kaitlyn Rudolf, Henry C. Hrdlicka, Peter Grevelding, Matthew Nankin

## Abstract

**Introduction:** Impaired balance-regulation after stroke put patients and therapists at risk for injury during rehabilitation. Body-weight-support systems (BWSSs) minimize this risk and allow patients to safely practice balance activities during therapy. Treadmill based balance perturbation systems with BWSSs are known to improve balance in patients with age or disease related impairments. However, these stationary systems are unable accommodate complex exercises requiring more freedom of movement.

**Objective:** To evaluate the impact of a new balance perturbation module, which is directly integrated to a track-mounted BWSS, has on patient balance after acute stroke.

**Design:** Unblinded quasi-randomized controlled pilot study.

**Setting:** Rehabilitation centered long-term acute care hospital.

**Participants:** Stroke rehabilitation inpatients with an admission Berg Balance Scale (BBS) assessment score ≥21/56.

**Interventions:** BWSS and BWSS with perturbation (BWSS-P) training was incorporated into the participants’ regular treatment. While both groups conducted the same balance and gait activities during their treatment sessions, the BWSS-P sessions included lateral and anterior/posterior resistive or assistive balance perturbations.

**Main outcome measures:** BBS and Activities-Specific Balance-Confidence (ABC) assessments were the main outcome measure collected. Institutional BBS data from fiscal-year 2018, prior to installation of the track mounted BWSS, was used as a historical standard-of-care (SOC) baseline.

**Results:** Improved post-intervention BBS and ABC assessment scores showed all participants benefited from therapy (p≤0.0438). The BBS percent-change of the BWSS-P [mean (SD) n] [66.95% (43.78%) 14] and BWSS control [53.29% (24.13%) 15] were greater than the SOC group [28.31% (17.25%) 30] (p≤0.0178), with no difference between BWSS groups (p=0.6669); ABC percent score-changes were also similar (p≥0.8036).

**Conclusions:** BWSS groups demonstrated similar BBS and ABC score improvements, indicating balance perturbations are not detrimental to post-acute stroke rehabilitation and are safe to use. This data provides strong rationale for conducting a larger follow-up study to further assess if this new perturbation system provides additional benefit to stroke patient balance during rehabilitation.

**Clinical Trial Registration:** NCT04919161

## Introduction

Each year, more than 795,000 people experience a stroke.^1^ Stroke, or cerebral vascular accident, is a neurological event that can lead to devastating physical and cognitive deficits, such as the inability to ambulate, impaired balance regulation, loss of coordination, and impaired communication.^2^ Due to the physical and cognitive deficits experienced following a stroke, many will require admission to an inpatient rehabilitation facility with the goal of maximizing their independence before returning to the home setting.^3^ Gait dysfunction is a common secondary impairment to stroke, usually requiring specific rehabilitative actions.^4^

Using motion analysis, patients after stroke are often observed navigating obstacles more conservatively and with abnormal gait patterns.^5^ This is likely associated with the loss of muscle-strength secondary to stroke, which could increase the risk of falling.^5^ Within six-months of discharge, falls occur in up to 70% of patients post-stroke, highlighting the importance of focusing on improving patients’ balance and gait during the early rehabilitation phase.^6^

It is estimated that over 90% of stroke survivors would report that the fear of falling negatively impacts their performance of daily living activities.^7^ Fear of falling has been shown to influence balance and gait control in older adults, supporting the theory that balance and gait should be taken into account during rehabilitative methods.^7^ These psychological factors are also strong predictors of falling compared to physical factors or the presence of pathology. Patient self-assessments can be important indicators of fall risk, as patients may better understand their capabilities/limitation than what the physical tests demonstrate.^8^

The ability to walk, stand, climb stairs, and other mobility-related functional tasks, are critical components of achieving functional independence. However, it is often difficult for post-stroke patients with balance impairments to safely practice balance and gait training without putting both therapists and patients at risk for injury. Incorporating robotic technologies to neurological rehabilitation can play a critical role in delivering safe and effective gait and balance therapy.^9^

The integration of body-weight support systems (BWSSs) following a stroke, spinal cord injury, or other neurological disorders has continued to expand over the last two decades, and the range of tools that therapists can use to treat these patients continues to grow.^10^ Using BWSSs to unload paretic lower limbs, patients with gait impairments can practice a higher repetition of steps in a safe, controlled manner. As the patient performs gait training, these systems support the patient’s body-weight, permitting patients with excessive weakness and poor coordination, to ambulate and perform more intensive therapy sessions sooner in their recovery, with minimal risk injurious fall. In addition to BWSSs, balance perturbation systems, which purposefully unbalance a patient so they can rehabilitate their postural control, have been used to improve gait and balance-control after stroke, or other age and disease related balance impairments.^11–16^

The goal of this study was to evaluate the efficacy of a recently developed, not yet reported, balance perturbation module for the ZeroG BWSS. This new balance perturbation training module is directly integrated into the BWSS and allows therapists to induce safe lateral, anterior, or posterior perturbations via a Wi-Fi-enabled handheld device. During both stationary and ambulatory activities, this system was used unbalance participants in order to train their balance-control and balance-reactions. The purpose of this pilot study was to determine if this newly developed BWSS balance perturbation system more effectively rehabilitates patient gait and balance after stroke than the standard BWSS protocol without perturbations.

## Methods

### Research Design

This was an unblinded quasi-randomized parallel controlled pilot study conducted at Gaylord Specialty Healthcare (Wallingford, CT, USA), a long-term acute care hospital. Before participant recruitment, the study was reviewed and approved by the hospital’s Institutional Review Board to ensure the study complied with the ethical-standards set by the Declaration of Helsinki and CONSORT 2010 (**Supplemental Materials**).^17^ The authors humbly admit that there was a delay in clinical trial registration for this pilot study, simply due to a lack of familiarity and awareness of the requirement for clinical trial registration and the definition of an applicable clinical trial.^18^ We are pleased to report this has been rectified and that retrospective clinical trial registration for this pilot study has been completed (NCT04919161).^19^

### Participants

All participants were admitted to the hospital under the inpatient stroke rehabilitation program after receiving a stroke diagnosis at a regional acute care hospital. Participant recruitment occurred over 12 months from October 2019 through September 2020. Patients admitted under the inpatient stroke rehabilitation program were evaluated by physical and occupational therapy within the first 72 hours of admission, at which point, an initial Berg Balance Scale (BBS) score was obtained as appropriate. To be considered, patients had to be classified as a “Moderate” fall risk, or better, shown by a BBS score of 21 or greater during their initial physical therapy evaluation. Patients who did not meet this inclusion criteria during their initial evaluation were able to screen-in at a later time pending a BBS reassessment. If the reassessment showed a sufficient functional improvement, and the patient had a discharge date of greater than two-weeks after the reassessment, the patient was recruited for the study.

In addition to meeting the BBS score criteria, participants needed to be 18 years of age or older, be able to understand and respond to simple verbal instructions in any language, and be able to tolerate and actively participate in at least three, 30 minute, weekly sessions in the BWSS. Patients were ineligible to participate if they did not meet one of these criteria or presented one or more of the exclusion criteria shown in **Table 1**.

**Table 1.** Exclusion Criteria for Study Participation.

|  |
| --- |
| Cognitive deficits that would disrupt the ability to provide informed consent |
| Berg Balance scale score less than 21 |
| Seizure |
| Spinal stabilization with use of Halo device |
| Uncontrolled hypo/hypertension |
| Unstable skin structures (i.e. skin grafts, chest tubes) |
| Unstable rib or lower extremity fractures |
| Osteoporosis |
| Active enteric infection control precautions |
| New limb amputations |
| Need for greater than 50% high flow oxygen |
| Bodyweight of more than 450 pounds (structural limitation of the BWSS) |

After providing informed consent, participants were assigned in an alternating fashion by the investigators to either the BWSS control or BWSS with perturbation (BWSS-P) group. Out of 50 patients approached for inclusion, 32 participants were enrolled, and 29 completed the study (**Figure 1**).

**Figure 1.**
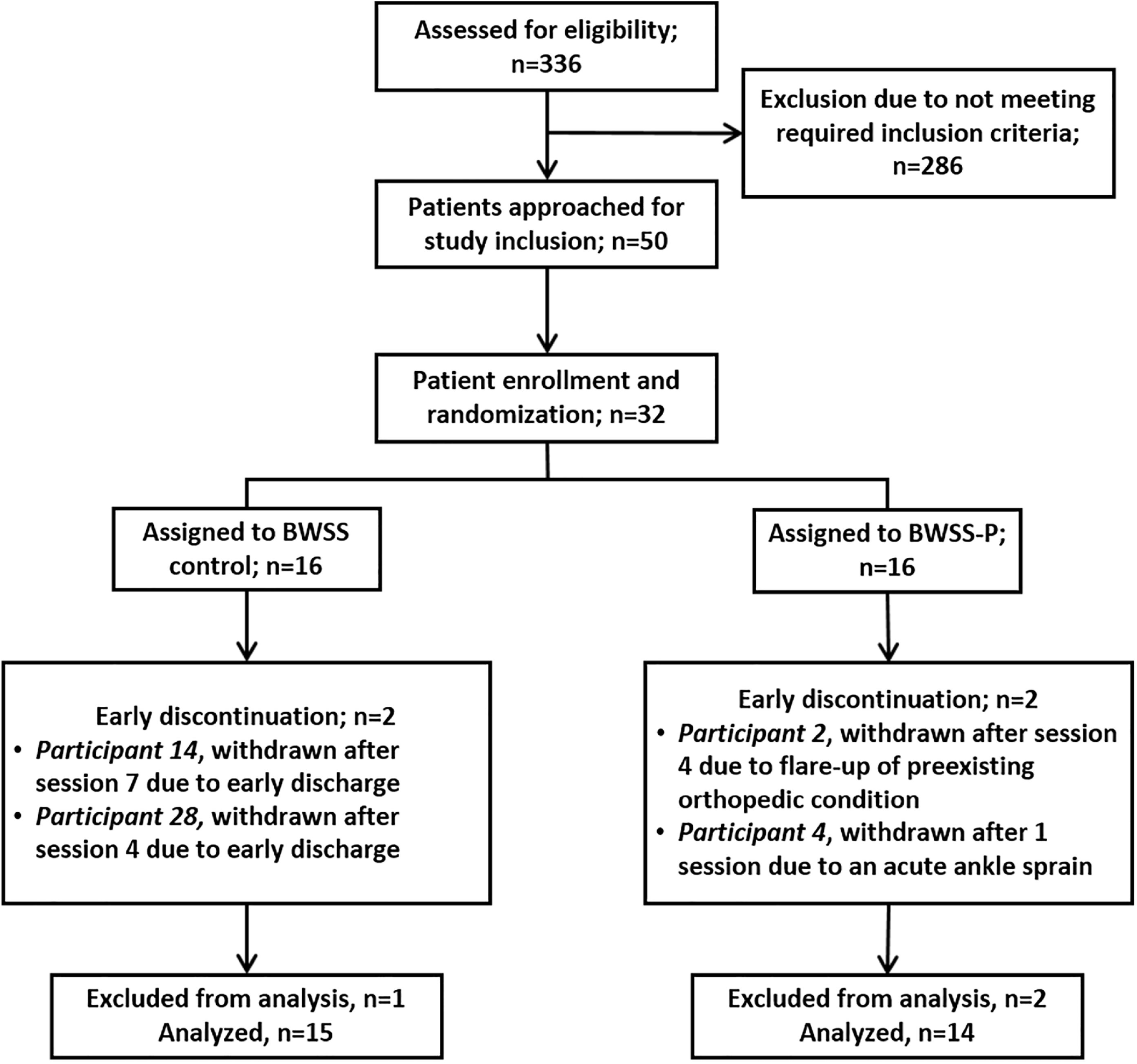
Participant Flow-cart. Of the approximately 336 patients admitted for stroke rehabilitation that were assessed for study eligibility, 50 were approached for study inclusion. Ultimately, 32 participants were consented, enrolled in the study, and alternately assigned to either the BWSS control or BWSS-P groups. During the study, 4 participants withdrew from the study early; 2 due to early discharge (after session 4 and 7 respectively), 1 due to a flare-up of a preexisting orthopedic condition after session 4, and 1 due to an acute ankle sprain after session 1. Data of the 2 participants who completed only 1 or 4 sessions were excluded from the data analysis.

Patients admitted for acute stroke rehabilitation typically receive 2 to 5 hours of skilled rehabilitative services 5 to 6 days per week, including physical, occupational, and speech therapies, and therapeutic recreation. All participants enrolled in this study were appropriate to receive this level of care.

### Instruments

The BBS and the Activities-Specific Balance Confidence (ABC) scale were the primary study endpoints. Both assessments have been validated for the stroke population and have high inter-rater reliability.^20,21^ The BBS is a standardized balance assessment that uses various balance tasks to objectively measure a person’s balance, and determine if a participant is a low, moderate, or high fall risk. The ABC Scale subjectively measures a person’s self-perceived balance-confidence to perform various tasks without losing balance or experiencing a sense of unsteadiness; it is based on a rating scale from 0% (no confidence) to 100% (completely confident).^8,21^

To identify eligible candidates for the study, chart reviews were conducted to collect the admission BBS scores of recently admitted stroke patients. The progression of patients who were disqualified from the study by just their admission BBS score were tracked through periodic chart reviews to determine if they had sufficiently improved to be re-considered for the study. During their regular treatment, a modified functional independence measure was used to assess each participant’s assistance needs while ambulating and undergoing toilet transfers (**Supplemental Materials**).^22^ A final chart review was conducted at the end of the study to collect participants’ BBS score and modified functional independence measures from their physical therapy discharge documentation. The ABC scale was administered pre and post-intervention by site investigators at the time of consent and immediately after the last intervention session.

### BWSS Equipment and Interventions

For this study, the BWSS used was the FDA listed ZeroG Gait and Balance System (Aretech, LLC, Ashburn, VA).^23^ We first introduced ZeroG to our institution in September 2019. Unlike some BWSSs, this device is mounted on an overhead track that follows patients as they ambulate.^23,24^ Like other BWSSs, this system is designed to unload the patient of up-to 200 pounds of their body weight while simultaneously protecting patients from falling. For this study, 10 pounds of participants’ body-weight, the system minimum required to engage the BWSS, was continuously displaced. If a patient were to fall, the system would detect the change, decelerate, and stop the descent after a set distance; the fall distance was set between 8 to 12 inches for the purpose of this study.

Unlike other BWSSs, a newly developed balance perturbation module known as the Training Responses in Postural rehabilitation or TRiP, is directly integrated to the ZeroG BWSS. This perturbation module is different than other systems as the balance perturbations are elicited directly through the BWSS and do not require a treadmill,^11–14^ tilt-table/shaking platform,^14,15^ or manual exertion by a therapist.^16^ Further, they can be induced during normal gait and balance exercises during therapy. The BWSS control group interventions consisted of various balance activities, including: marching, side-stepping, retro-ambulation, step-taps, and step-ups. The BWSS control group also practiced various gait tasks, including: ambulation over the ground, going up and down stairs, and performing sit-to-stand transitions. The BWSS with perturbation (BWSS-P) intervention group performed the same activities as the control group, with just the addition of lateral, anterior, and posterior perturbations. Assistive devices and equipment were used during intervention sessions as recommended by the participant’s primary therapist, including: canes, rolling walkers, hemi-walkers, and ankle-foot-orthoses (AFO), ankle support braces, and upper extremity slings.

Therapist administered perturbations using a Wi-Fi-enabled handheld device linked to the BWSS and these consisted of a sudden and brief assistive or resistive force in the desired direction. Lateral perturbations were issued while participants were in a static stance, while anterior and posterior perturbations were issued during ambulation; eight perturbations, two in each direction, were completed each session.

All participants started at perturbation level “one” and progressed up to a maximum perturbation level of “ten” through the course of the study. The amount of force exerted at each perturbation level is pre-set by the manufacturer. The perturbation level (i.e. intensity or force) used each session was based on the participant’s progress and observational analysis made by the therapist of the participants’ response to the perturbation level. If the participant was able to tolerate the initial perturbation level without exhibiting a balance reaction, the perturbation level was incrementally increased until an appropriate balance reaction was exhibited. If a participant was unable to recover and elicited a fall response in the system, the perturbation level was decreased by one level to ensure patient safety, and the exercise repeated to reinforce the exercise mechanics and participant confidence. The highest perturbation level achieved was recorded after each session is what is reported.

Participants in both study groups received a total of eight treatment sessions over two weeks. As necessary, participants received up to two sessions in one day to ensure they completed the required eight sessions before discharge. These sessions were incorporated into the participants’ regular care. At our institution, treatment sessions are broken into 30 minute blocks. This time includes patient transportation, equipment set-up, and in the case of this study, donning the BWSS harness. On average, participants received 20 minutes of active time in the BWSS for each 30 minute treatment block. All sessions were analyzed equally despite the length of time in the BWSS.

### Data Analysis

Data was analyzed using GraphPad Prism version 9.0.0 (GraphPad Software, San Diego, CA). To compare the observed proportion of males and females in the BWSS groups, a Binomial Test and Fisher’s exact test were used. The 95% CIs reported for the proportion of males and females in the BWSS-P group were calculated using the Wilson-Brown Method.

BBS and ABC measurements changes between the pre- and post-intervention were compared directly, as well as between groups. The degree of change made by each individual was shown by calculating the *percent change*:

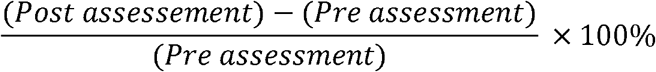

BBS data of stroke rehabilitation patients from fiscal year 2018 served as a historical standard of care (SOC) baseline control. The SOC data was sorted to consist of patients with initial BBS scores of 21 or greater and who were admitted and discharged before the launch of the institution’s BWSS in September 2018. This resulted in the inclusion of retrospective BBS data from 30 patients.

Shapiro-Wilk testing was first used to test for normality; if one or more of the data-sets in the group failed (p<0.05), nonparametric tests were used going forward. For hypothesis testing between two groups, unpaired or paired two-tailed Student’s t-test were conducted as appropriate. When indicated by an F-test for variance (p<0.05), Welch’s correction was applied for unequal standard deviations between groups.

When comparing three or more groups, if one or more groups were abnormally distributed, non-parametric Kruskal-Wallis analysis of variation (ANOVA) test and Dunn’s multiple comparison test for statistical hypothesis testing were used. When normally distributed, an Ordinary one-way ANOVA with a Tukey’s multiple comparisons test for statistical hypothesis testing was used. If Brown-Forsythe’s test for variance indicated the variance of the groups were significantly different (p<0.05), a Brown-Forsythe correction was applied and Dunnett’s T3 multiple comparisons test for statistical hypothesis testing was used instead.

For data represented as a box-plot, each box represents the median and the 25% and 75% quartiles respectively. The whiskers extend 1.5 and -1.5 of the interquartile range respectively, triangle symbols reflect data-points beyond the 1.5 interquartile ranges, and the ‘+’ symbol represents the arithmetic mean.

## Results

Of the 29 participants who completed the treatment course, 15 were alternately assigned to the BWSS control group and 14 were alternately assigned to the BWSS-P group. There were 13 males and 2 females in the BWSS group, and 10 males and 4 females in the BWSS-P group **(Table 2)**. One participant in the BWSS control group did not complete the eighth and final session due to an early discharge; the data from the seven completed sessions were included in the analysis. Compared to the control group, the BWSS-P group was similarly aged (p=0.9427) and had similar proportions of males and females (p=0.3898). The baseline characteristics of the study groups were not significantly different (**Table 2**).

**Table 2.** Participant Demographics.

|  | **BWSS Control** | **BWSS-P** |
| --- | --- | --- |
| Number of participants (n) | 15 | 14 |
| Cohort age, mean (SD) | 57.8 (13.0) | 57.5 (14.2) |
| Male n (%) | 13 (86.7%) | 10 (71.4%) |
| Male age, mean (SD), range, years | 57.5 (12.7), 37 | 57.4 (11.3), 31 |
| Female n (%) | 2 (13.3%) | 4 (28.6%) |
| Female age, mean (SD), range, years | 60.5 (20.5), 29 | 57.8 (22.2), 50 |
| Fisher's exact test (two-sided)<sup>†</sup> | p=0.3898 |  |
| Two-tailed Binomial test<sup>†</sup> | p=0.2587 |  |
| Confidence Intervals | Upper bound | Lower bound |
| Wilson-Brown 95% CI of proportion of BWSS-P Males | 45.35 | 88.28 |
| Wilson-Brown 95% CI of the proportion of BWSS-P Females | 11.72 | 54.65 |

*SD, standard deviation; CI, confidence interval; BWSS, body-weight support system; BWSS-P, body-weight support system with perturbations*
<sup>†</sup>Fisher's exact test and a binomial test were used to compare the proportion of males and females between the BWSS groups.

Throughout the study, most participants tolerated the BWSS induced perturbations well, however two of the 32 original participants enrolled in the study did not complete all eight therapy sessions due to injury. One participant experienced an unexpected flare-up of a preexisting chronic orthopedic condition unrelated to the BWSS perturbation module. A second participant suffered an acute ankle sprain during ambulation in the BWSS. The nature of this injury was deemed likely a combination of the BWSS perturbation module and ankle instability secondary to the participant’s stroke. A third participant also withdrew early from study due to an early discharge (**Figure 1**).

From the BWSS perturbation module, the highest perturbation level achieved for each patient, each session, was recorded. Although the final perturbation level achieved by the final session varied, all participants showed significant improvement by the end of the study (p<0.0001, **Figure 2A**). These data can then be divided into three categories. First, the “low responders” showed early perturbation level progression but plateaued, peaking at perturbation levels 4 to 5 (**Figure 2B**). The “moderate responders” showed steady progress throughout the study, peaking between perturbation levels 6 to 8 (**Figure 2C**). The “high responders” rapidly progressed through the BWSS-P levels, peaking between BWSS-P levels 9 to 10 (**Figure 2D**).

**Figure 2.**
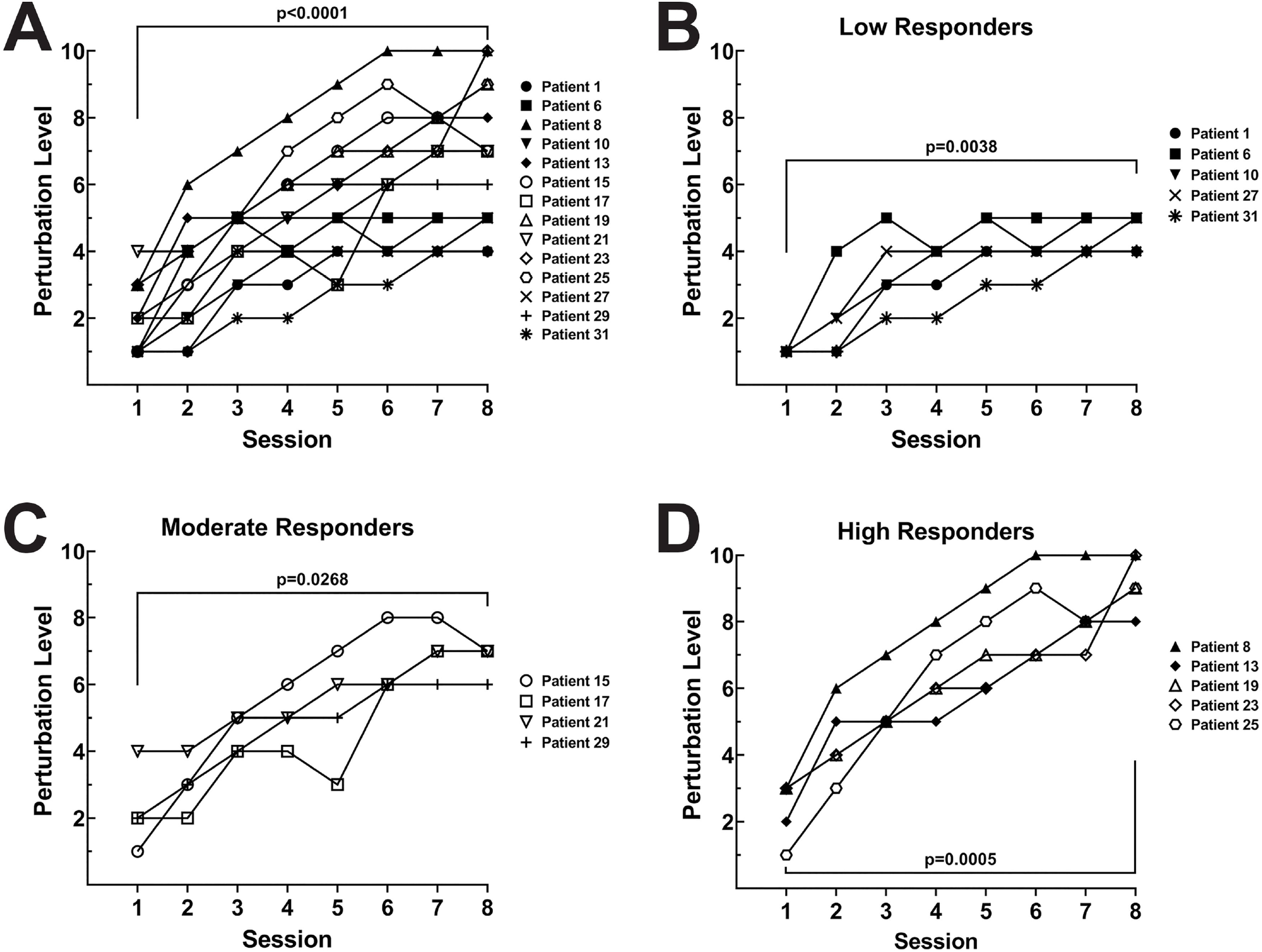
Perturbation level progression. From the BWSS, the highest perturbation level achieved was recorded for each participant, after each therapy session. Although the final perturbation level achieved was variable, each participant who completed the study successfully increased their perturbation level between the first and last therapy session **(A)**. The perturbation level progression for the participants that completed the study could be broken down into three categories, Low Responders **(B)**, Moderate Responders **(C)**, and High Responders **(D)**.

The BBS assessment data showed that individuals in all three groups, SOC, BWSS control, and BWSS-P, improved pre-to post-intervention (**Figure 3A, Table 3**, SOC p=0.0001, BWSS p=0.0025, BWSS-P p=0.0001). However, comparison of the pre- and post-intervention scores of the three cohorts revealed that the mean baseline and mean outcome measurements of the three groups were similar.

**Figure 3.**
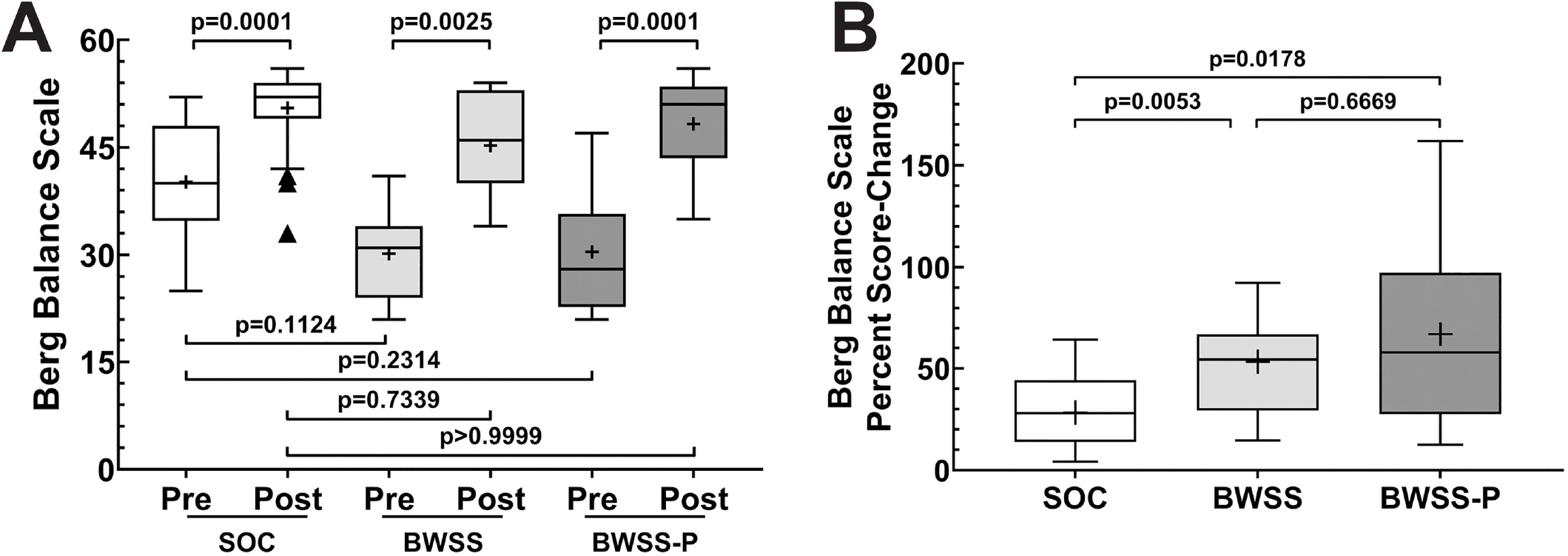
Berg Balance Scale assessment. Participant’s pre and post-intervention BBS assessment scores were used to track their improvement/response to the therapy. In addition to the BWSS control and BWSS-P protocols, facility data from fiscal year 2018 prior to the implementation of the BWSS, served as a historical standard of care (SOC) control. We examined the raw scores in aggregate **(A)**. We also normalized the BBS score data by calculating the percent change for each participant to show the magnitude of change between pre and post-intervention **(B)**. The box-plot represent the median and the 25% and 75% quartiles respectively. The whiskers extend 1.5 and -1.5 of the interquartile range respectively; triangle symbols reflect data-points beyond the 1.5 interquartile ranges; “+” symbols represents the mean; SOC n=30, BWSS control n=14-15, BWSS-P n=13-14.

**Table 3.** Summary of Berg Balance Scale Assessments.

|  | **SOC** |  | **BWSS** |  | **BWSS-P** |  |
| --- | --- | --- | --- | --- | --- | --- |
| **Parameters** | **Pre** | **Post** | **Pre** | **Post** | **Pre** | **Post** |
| Number of individuals | n = 30 |  | n = 15 |  | n = 14 |  |
| Mean | 40.20 | 50.50 | 30.20 | 45.27 | 30.43 | 48.29 |
| Standard Deviation | 7.66 | 5.41 | 6.41 | 6.67 | 7.97 | 6.94 |
| Minimum | 25 | 33 | 21 | 34 | 21 | 35 |
| Median | 40 | 52 | 31 | 46 | 28 | 51 |
| Maximum | 52 | 56 | 41 | 54 | 47 | 56 |
| Percent Change, Mean (SD) (%) | 28.31(17.25) |  | 53.29 (24.13) |  | 66.95 (43.78) |  |

*SOC, Standard of Care; BWSS, body-weight support system; BWSS-P, body-weight support system with perturbations; SD, Standard Deviation.*
Percent change was calculated as [(post score – pre score)/(pre score) x 100%]

**Figure 4.**
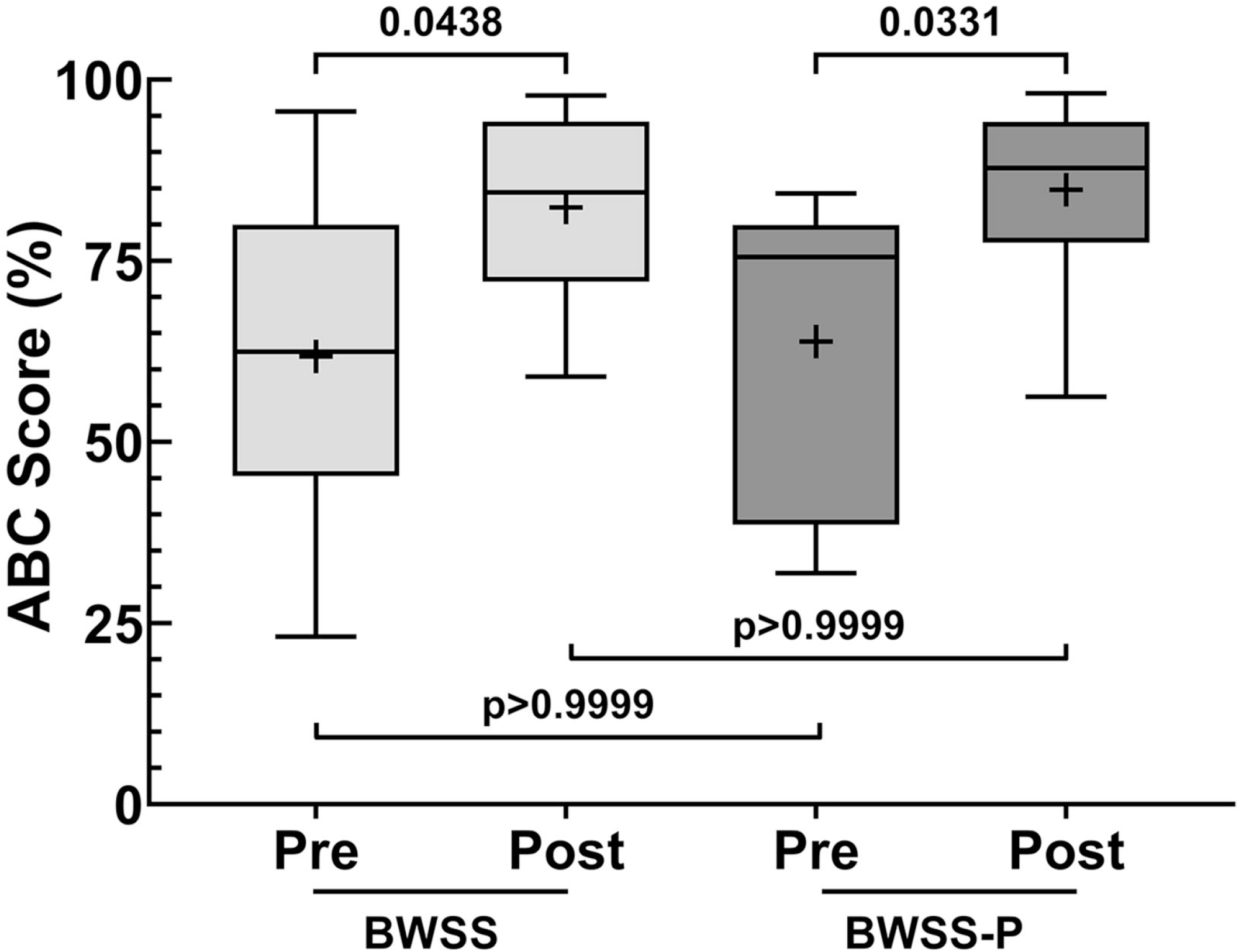
Activities Balance Confidence (ABC) scale assessment. The ABC assessment was also given to participant’s pre and post-intervention to gauge their confidence in performing daily tasks. The box-plot represent the median and the 25% and 75% quartiles respectively. The whiskers extend 1.5 and -1.5 of the interquartile range respectively; “+” symbols represents the mean; BWSS control n=14-15, BWSS-P n=13-14.

Observing this, we assessed the degree of change for each individual by calculating the percent change (**Figure 3B**). This analysis showed that, while the mean pre- and post-assessment scores were similar between groups, there was a greater [mean (SD), n] percent change in the BWSS-P group [67.0% (43.8%), 14] compared to the SOC group [28.3% (17.3%), 30] (p=0.0178). The percent change of the BWSS control group [53.3% (24.1%), 15] was also found to be greater than the SOC group (p=0.0053). While the percent change of the BWSS-P group was slightly greater than the BWSS control group, it was not significantly different (p=0.6669).

Modified functional independence measure scores (**Supplemental Materials**) were used to assess each participant’s functional independence during ambulation and toilet transfers. The [mean (SD)] ambulation assistance score increased in both the BWSS control [4.36 (1.03) to 7.80 (1.20)] and BWSS-P treatment [4.75 (0.83) to 8.64 (0.93)] groups (p<0.0001). Similarly, the [mean (SD)] toilet transfer score increased in both the BWSS control [4.30 (0.59) to 7.70(1.16)] and BWSS-P treatment [4.89 (0.79) to 8.39 (1.04)] groups (p<0.0001) (**Supplemental Materials**). The pre- and post-intervention modified functional independence measure scores of the BWSS groups were virtually identical (p>0.9999).

Participant self-confidence in performing daily tasks was evaluated using the ABC scale. Similar to the BBS, participants reported increased confidence in their ability to perform daily tasks after the intervention (BWSS p=0.0438, BWSS-P p=0.0331). However, the pre- and post-intervention ABC scores were virtually identical between BWSS groups (p>0.9999) **(Figure 3)**.

## Discussion

To improve patient balance after an acute stroke in the long-term acute care hospital setting, we evaluated the effectiveness of a new, BWSS integrated, balance perturbation training module. This module induces controlled balance perturbations during gait and balance exercises without a treadmill or other equipment. Participants in both BWSS groups demonstrated similar, significant improvement in their BBS, ABC assessment, and ambulation and toileting transfer scores. At a minimum, this indicates the BWSS-P protocol is not detrimental and may benefit post-acute stroke rehabilitation. With fiscal year 2018 BBS data serving as a historical SOC control, both BWSS groups displayed greater BBS percent score changes than the SOC group. This data supports the overall conclusion that this new BWSS balance perturbation module may help to improve patient balance after acute stroke when following a prescribed treatment and rehabilitation plan.

Conventional balance perturbation training, including modified treadmills,^11–14^ tilt-tables,^14,15^ or external force provided by the therapist directly,^16^ pose an injury risk to the therapist and the patient. Additionally, if the patient were to experience an injurious fall during treatment, it may further contribute to their fear of falling after stroke. While incorporation of BWSSs over treadmills decreases the injury risk, this is not representative of functional ambulation in the patients’ home or community environment.^25,26^ Further, these strategies are stationary and limit the types of activities and exercises that can be performed during balance perturbation (i.e. navigating a turn). Systems such as these may also limit the participation of some patients who would otherwise benefit from balance perturbation training, such as patients uncomfortable or unable to ambulate on a treadmill.

Therapists also have the option of inducing balance perturbations by manually exerting an external force (i.e. pulling or pushing the patient) while a patient is in a BWSS. While more accessible than using specialized equipment, the application of force by the therapist and amount of perturbation experienced by the patient is subjective, and could be hard to control and/or replicate consistently. Integration of the balance perturbation module to BWSSs described here resolves many of these issues, including: allowing for freedom of movement to perform most gait and balance exercises in a dynamic environment; increasing the accessibility to eligible patients; and performing perturbations in a consistent, repeatable, and quantitative manner while optimizing therapist and patient safety.

In this pilot study, it was difficult to attribute improvements to the BWSS perturbation module alone, as both BWSS groups showed similar BBS score improvements. Although the mean scores were not significantly different, the variability of the initial BBS scores of the BWSS study groups may have limited our ability to accurately determine the impact of the perturbation module. This variability, in part, is reflective of the diverse patient population that was recruited; any qualifying stroke inpatient with a BBS of 21 or greater were approached. While this was addressed in the data analysis by calculating the percent change for each participant, we could improve this variability in future studies in one of several ways.

First we could stratify our data analysis and compare the amount of change/improvement at different admission BBS scores to better refine what populations benefit the most from this treatment. We could also include an upper BBS score to our inclusion criteria. For example, for acute stroke a cut-off of 45/56 has been used to describe a normal functional ability post-stroke.^27^ Lastly, a matched-control method could be implemented to ensure the same range of initial BBS score are represented in the BWSS groups. In any case, a larger population will be required in future studies to achieve the appropriate power needed to observe the impact of the BWSS perturbation module.

Variability in the timing of the post-intervention BBS assessments may have also contributed to the lack of significant difference between BWSS groups. The post-intervention BBS scores were obtained by the participants’ primary physical therapist at the time of their discharge. Most participants had discharge dates close to the last session of the study intervention. However, this does not account for progress the participant may have made after the last session leading up to their discharge date, especially if there was an unexpected delay in their discharge. To address this in future studies, we propose delivering a separate BBS assessment within 48 hours of the last session, if the participant’s discharge assessment is not collected during that time.

Most participants completed the study-related sessions over a two-week period, however, this study was partly conducted during the early months of the COVID-19 pandemic (03/2020-08/2020). This environment may have shortened the amount of time eligible patients were willing to spend in an inpatient setting if they were able to safely navigate the home environment with assistance. Due to this, many patients who met the inclusion criteria for the study did not remain an inpatient long enough to receive the required eight sessions. As a further consequence of expedited discharge dates due to COVID-19, 40% of participants, at least once, needed to receive two sessions per day to complete all eight sessions; in one case, a patient was discharged before they were able to complete their last treatment session. It is unclear if the increased intensity positively or negatively contributed to the rate of progress. To address the possibility of irregular lengths of stay in the future, we will evaluate and compare the dose-response relationship of the perturbation system over two to six sessions, as well as the total time in the system. This would open-up the recruitment pool to eligible participants with a shorter length of stay and allow us to refine the optimal dosing. These studies could also investigate how many sessions per day and per week is most effective at improving balance control, reaction, and confidence.

The study was also strengthened by the quasi-randomized controlled design and low participant dropout rate (3/32; 9.4%). With 52 years of combined experience, the study was further reinforced by the advanced specialty and board certifications of the treating investigators.

This pilot study provides a strong foundation to examine the impact of the BWSS-P protocol on a larger population. It also provides the preliminary data necessary to conduct a power analysis of the effect size and the population needed going forward. A larger multi-site study would also allow us to better generalize the effectiveness of BWSS-P on other outcomes such as discharge destination and falls after discharge. Furthermore, we could also expand our scope and examine how other variables, such as stroke location or other compounding diagnoses, impact patient progress and response to balance perturbation training using this system. Incorporating additional dynamic gait assessments that more closely resembles functional movement patterns and/or reactive-balance specific outcome-measures, such as the Dynamic Gait Index^28^ or Functional Gait Assessment,^29^ to future studies may assist us in better understanding the full implications of this new balance perturbation module.

## Conclusion

In conclusion, this study has multiple implications for clinical practice in the inpatient rehabilitation setting. The BWSS-P positively impacted the balance performance of a subset of stroke inpatients who scored greater than or equal to 21 on their BBS assessment. Not only did the BWSS-P improve participant balance and decrease their fall risk compared to the SOC, it improved participants’ overall confidence and reduced their fear of falling similar to using the BWSS alone. As this is the first study to investigate the utility of a track-mounted BWSS integrated balance perturbation module such as ZeroG TRiP system, there are a number opportunities for continued research and development in this area going forward.

## Data Availability

Copies of the study protocol will be provided upon request. Requests for copies of the de-identified research data will be considered on a situational basis.

## Declarations

### Patient Consent

Before participant recruitment, the study was reviewed and approved by the hospital’s Institutional Review Board to ensure the study complied with the ethical-standards set by the Declaration of Helsinki. Written informed consent was delivered and collected from each participant before study activities were conducted.

### Protocol and Data Sharing

Copies of the detailed treatment protocol is available upon request. Requests for copies of the de-identified datasets will be considered upon request as appropriate.

## Acknowledgments

The authors would like to acknowledge Dr. Richard Feinn of Quinnipiac University for their assistance with the preliminary data analysis. We also want to acknowledge all of our colleagues who provided their careful and thoughtful review of the manuscript.

## Abbreviations

BBS: Berg Balance Scale
ABC: Activities-Specific Balance-Confidence
SOC: Standard of Care
BWSS: body-weight support system
BWSS-P: body-weight support system with perturbations
TRiP: Training Responses in Postural-Rehabilitation

