## Supplemental Materials for "Inducing Body-Weight Supported Postural Perturbations during Gait and Balance Exercises to Improve Balance after Stroke – A Pilot Study"

**Supplemental Table 1. Modified functional independence measure definitions and criteria**

| Score | Descriptor [a, b] | Definition |
|---|---|---|
| 1 | Dependent (D) | Dependent mobility; subject/patient providing less than 25% of the work |
| 2 | Maximal Assistance (MAX) | Subject/patient performs 25 to 49% of the work |
| 3 | Moderate Assistance (MOD) | Subject/patient performs 50 to 74% of the work |
| 4 | Minimal Assistance (MIN) | Subject/patient performs 75 to 100% of the work |
| 5 | Contact Guard Assist (CG) | Subject/patient requires light hands on assistance for balance but no physical lifting is required |
| 6 | Close Supervision (CS) | Subject/patient requires the therapist to be close by in case the patient experiences a loss of balance, but does not provide physical or hands on assistance |
| 7 | Supervision (S) | During supervision the therapist is proving supervision at more than an arm length away. |
| 8 | Distant Supervision (DS) | This is "intermittent supervision." The therapist does not have to be in the room. |
| 9 | Modified Independence (MOD I) | The subject/patient is independent WITH use of adaptive device, techniques, or increased time. |
| 10 | Independent (I) | The subject/patient is independent WITHOUT use of adaptive device, techniques, or increased time. |

[a] During treatment, the abbreviated descriptors were recorded in the patient's chart, but they were codified using the associated score shown to better facilitate statistical analysis.
[b] Instances when two descriptors were recorded, the average score of the listed descriptors were used (i.e. CS/CG = 5.5 and CG/MIN = 4.5).

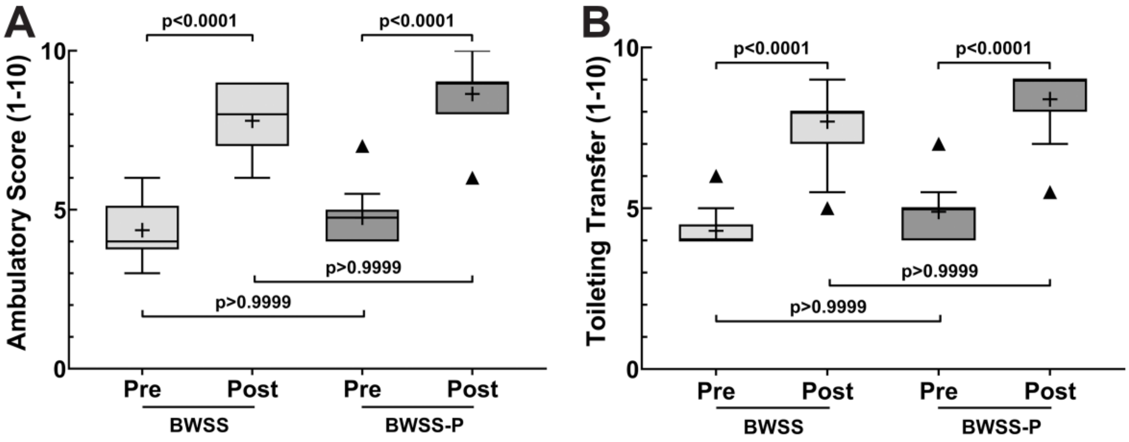


**Supplemental Figure 1. Ambulation and Toileting Transfer Assistance mFIM Scores.** In addition to evaluating patient functional outcomes by the BBS, we also measured the level of assistance participants required during ambulation **(A)** and toilet-transfer **(B)**. These were measured using the modified FIMS scale shown in **Supplemental Table 1**. The box-plot represent the median and the 25% and 75% quartiles respectively. The whiskers extend -1.5 and 1.5 of the interquartile range respectively; triangles reflect data-point values outside the interquartile range; + represents the mean; BWSS n=14-15, BWSS-P n=13-14

### CONSORT 2010 checklist of information to include when reporting a randomised trial*

| Section/Topic | Item No | Checklist item | Reported on page No |
|---|---|---|---|
| **Title and abstract** | | | |
| | 1a | Identification as a randomised trial in the title | n/a |
| | 1b | Structured summary of trial design, methods, results, and conclusions (for specific guidance see CONSORT for abstracts) | 1-2 |
| **Introduction** | | | |
| Background and objectives | 2a | Scientific background and explanation of rationale | 3-4 |
| | 2b | Specific objectives or hypotheses | 4 |
| **Methods** | | | |
| Trial design | 3a | Description of trial design (such as parallel, factorial) including allocation ratio | 5 |
| | 3b | Important changes to methods after trial commencement (such as eligibility criteria), with reasons | 15 |
| Participants | 4a | Eligibility criteria for participants | 5-6, Table 1 |
| | 4b | Settings and locations where the data were collected | 5 |
| Interventions | 5 | The interventions for each group with sufficient details to allow replication, including how and when they were actually administered | 7-8 |
| Outcomes | 6a | Completely defined pre-specified primary and secondary outcome measures, including how and when they were assessed | 6-7 |
| | 6b | Any changes to trial outcomes after the trial commenced, with reasons | 15 |
| Sample size | 7a | How sample size was determined | 6 |
| | 7b | When applicable, explanation of any interim analyses and stopping guidelines | n/a |
| Randomisation: | | | |
| Sequence generation | 8a | Method used to generate the random allocation sequence | 6 |
| | 8b | Type of randomisation; details of any restriction (such as blocking and block size) | 6 |
| Allocation concealment mechanism | 9 | Mechanism used to implement the random allocation sequence (such as sequentially numbered containers), describing any steps taken to conceal the sequence until interventions were assigned | 6 |
| Implementation | 10 | Who generated the random allocation sequence, who enrolled participants, and who assigned participants to interventions | 6 |
| Blinding | 11a | If done, who was blinded after assignment to interventions (for example, participants, care providers, those assessing outcomes) and how | n/a |
| | 11b | If relevant, description of the similarity of interventions | 7-8 |

| | | | |
|---|---|---|---|
| Statistical methods | 12a | Statistical methods used to compare groups for primary and secondary outcomes | 9-10 |
| | 12b | Methods for additional analyses, such as subgroup analyses and adjusted analyses | 9-10 |
| **Results** | | | |
| Participant flow (a diagram is strongly recommended) | 13a | For each group, the numbers of participants who were randomly assigned, received intended treatment, and were analysed for the primary outcome | 10-11, Figure 1 |
| | 13b | For each group, losses and exclusions after randomisation, together with reasons | 11, Figure 1 |
| Recruitment | 14a | Dates defining the periods of recruitment and follow-up | 5, 15 |
| | 14b | Why the trial ended or was stopped | n/a |
| Baseline data | 15 | A table showing baseline demographic and clinical characteristics for each group | Table 2 |
| Numbers analysed | 16 | For each group, number of participants (denominator) included in each analysis and whether the analysis was by original assigned groups | 10-11, Figure 1, Figure legends |
| Outcomes and estimation | 17a | For each primary and secondary outcome, results for each group, and the estimated effect size and its precision (such as 95% confidence interval) | 10-12, Table 3 & Figure 2-4 |
| | 17b | For binary outcomes, presentation of both absolute and relative effect sizes is recommended | 10-12, Table 3 & Figure 2-4 |
| Ancillary analyses | 18 | Results of any other analyses performed, including subgroup analyses and adjusted analyses, distinguishing pre-specified from exploratory | 9, Table 3 & Figure 2-4 |
| Harms | 19 | All important harms or unintended effects in each group (for specific guidance see CONSORT for harms) | 11 |
| **Discussion** | | | |
| Limitations | 20 | Trial limitations, addressing sources of potential bias, imprecision, and, if relevant, multiplicity of analyses | 13-16 |
| Generalisability | 21 | Generalisability (external validity, applicability) of the trial findings | 13-16 |
| Interpretation | 22 | Interpretation consistent with results, balancing benefits and harms, and considering other relevant evidence | 13-16 |
| **Other information** | | | |
| Registration | 23 | Registration number and name of trial registry | Currently pending retrospective registration (p.5) |
| Protocol | 24 | Where the full trial protocol can be accessed, if available | 17 |
| Funding | 25 | Sources of funding and other support (such as supply of drugs), role of funders | 17 |
